# Modality Fusion of MRI and Clinical Data for Glioma Tumour Grading

**DOI:** 10.64898/2026.04.20.26351308

**Authors:** Reyhaneh Kheirbakhsh, Prateek Mathur, Aonghus Lawlor

**Affiliations:** Insight Research Ireland Centre for Data Analytics; School of Computer Science, University College Dublin, Dublin, Ireland

**Keywords:** Modality Fusion, Deep Learning, Glioma Grading

## Abstract

Multimodal machine learning leverages complementary information from diverse data sources and has shown strong promise in medical imaging, where multimodal data is critical for clinical decision making. In glioma grading, integrating MRI modalities with clinical data can improve diagnostic accuracy, yet systematic comparisons of fusion strategies remain limited. This study evaluates early, intermediate, and late fusion approaches, addressing the question: How does the inclusion of clinical data alongside MRI modalities influence grading performance? To assess modality contributions, we design adaptable fusion layers and employ interpretability techniques, including attention-based analysis. Our results show that incorporating clinical data consistently outperforms unimodal and MRI-only baselines, with intermediate fusion yielding the most reliable gains. Beyond accuracy, the framework reveals how MRI and clinical features jointly shape predictions, underscoring the importance of both fusion design and interpretability for clinical adoption.

## 1 Introduction

In recent years, multimodal machine learning (ML) has played an increasingly important role in healthcare by integrating diverse data modalities such as magnetic resonance imaging (MRI), histopathology, clinical data, and genomic profiles [1]. This growing interest is driven by both the availability of heterogeneous biomedical data and the recognition that combining complementary modalities enables a more comprehensive understanding of complex medical conditions [2]. The central value of multimodality lies in complementarity [3], where each modality contributes unique information. However, a key challenge is modality fusion, which involves developing strategies to effectively represent multimodal data while exploiting its complementarity. This is particularly difficult due to the heterogeneous nature of biomedical data [4].

Early and accurate detection is paramount in cancer diagnosis for significantly improving patient outcomes. This critical need is profoundly exemplified in neuro-oncology by gliomas, a heterogeneous group of primary brain tumours classified by the WHO into grades 1-4 based on histopathological and molecular characteristics. Accurate grading is essential, as treatment strategies and prognosis vary substantially across grades [5]. Although MRI is the standard for non-invasive assessment, it often lacks the specificity to capture full tumour heterogeneity. Integrating MRI with complementary data, such as clinical information, offers the potential to improve diagnostic precision [6]. Consequently, there is huge potential for multimodal ML to meet this challenge by providing more precise and rapid diagnostic capabilities.

While multimodal ML approaches for glioma grading exist [7,8,9] two key challenges remain: identifying the most effective strategy for integrating imaging and non-imaging data, and quantifying the contribution of each modality to performance. To address these gaps, we propose a multimodal fusion framework that leverages both MRI and clinical data, systematically evaluating early, intermediate, and late fusion. We also address interpretability through a keyless attention-based fusion [10] block. Our study addresses three key questions: (1) How does the inclusion of clinical data alongside MRI modalities influence grading? (2) How does intermediate fusion compare to early and late fusion in terms of predictive power? (3) Which modalities contribute most significantly to the overall performance of the multimodal model?

## 2 Related Work

The standard taxonomy for fusion level in multimodal ML [11] includes early fusion, which integrates data at the input level; late fusion, which aggregates outputs from separate models; and intermediate fusion, which combines modality-specific features during the learning process to allow interaction (fig. 1). The choice of fusion level often depends on the degree of correlation between modalities [12].

**Fig. 1:**
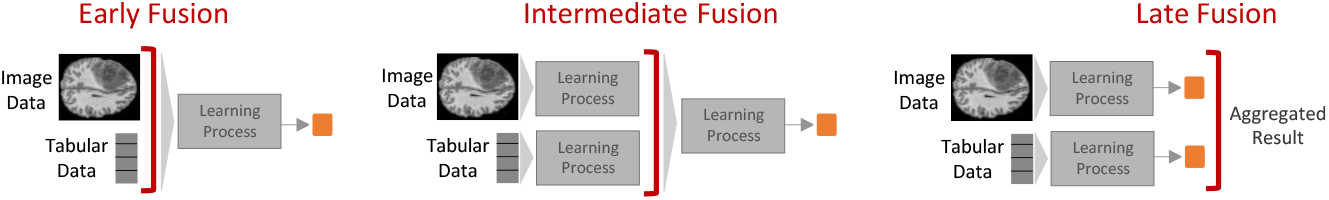
Overview of different levels of fusion for image and tabular data

Recent research has moved beyond simple concatenation to more expressive fusion mechanisms. Methods like high-order attention [13], dual attention, cross-attention, and transformer-based modules [14,15] are used to align multimodal information and adaptively weight modalities instead of treating them equally. This approach has led to significant performance gains, especially with heterogeneous healthcare data. In healthcare, multimodal approaches integrating image and non-image data show strong potential [1]. Examples include attention-based fusion for pan-cancer survival prediction [16] and Alzheimer’s diagnosis [17], demonstrating how complementary data can improve predictive performance.

Specific to glioma diagnosis, most studies focus on fusing multiple MRI modalities such as T1, T1c, T2, and FLAIR. For example, Guo et al. [18] developed a multimodal MRI model employing DenseNet for modality-specific feature extraction, followed by a linear weighted late fusion strategy to combine predictions. Guo et al. [8] introduced a graph convolutional network to integrate spatial and contextual MRI features. Beyond MRI, some studies have explored incorporating additional modalities. Hsu et al. [19], for instance, combined pathology images and clinical data with MRI using a late fusion approach, relying primar-ily on whole slide imaging (WSI)-based predictions and leveraging MRI-derived outputs as auxiliary guidance when confidence was low. In another work, Usuzaki et al. [9] propose a variable Vision Transformer (vViT), a self-attention–based model designed for glioma grading that integrates demographic data, radiomic features, and multi-sequence MRI.

A growing body of research emphasises not only improving predictive accuracy but also enhancing interpretability by identifying which modalities drive model decisions. For instance, Pathomic Fusion [7] integrates histology and genomic features for glioma survival prediction, modelling cross-modal interactions via Kronecker products and refining contributions through gating-based attention. Jeong et al. [20] employed an attention-based fusion block with ablation studies to assess the impact of missing modalities, while Byeon et al. [6] applied attention visualisation and relevance propagation to highlight focus regions and quantify modality contributions. Collectively, these works demonstrate how attention mechanisms and ablation strategies can enhance the transparency of multimodal fusion models, a key requirement for clinical applicability.

Despite these advances, prior work has largely focused on specific fusion methods, with limited attention to interpreting the contributions of individual modalities. A systematic comparison of early, intermediate, and late fusion under identical conditions, coupled with interpretability, remains underexplored. This gap motivates our framework, which evaluates different fusion levels while providing transparency into how individual modalities influence glioma grading.

## 3 Methodology

To evaluate the impact of integrating clinical data (age and sex) with MRI modalities (T1, T1c, T2, and FLAIR) across different fusion levels for glioma grading, we designed a framework encompassing five fusion strategies: two early fusion variants, two intermediate fusion variants based on feature concatenation, and one late fusion approach using majority voting and probability averaging. The contribution of MRI and clinical features is assessed through two complementary analyses. First, a modality ablation study compares performance with and without clinical data. Second, we introduce an attention-based fusion block to replace simple concatenation, enabling the computation of modality-specific contribution scores that quantitatively reflect the role of each modality in the classification process.

### 3.1 Fusion Strategies

The fusion architectures in our framework are illustrated in fig. 2. They are referred to as Early Raw Features (ERF), Early Learned Features (ELF), Intermediate Separate Features (ISF), Intermediate Merged Features (IMF), Late Probability Averaging (LPA), and Late Majority Voting (LMV).

**Fig. 2:**
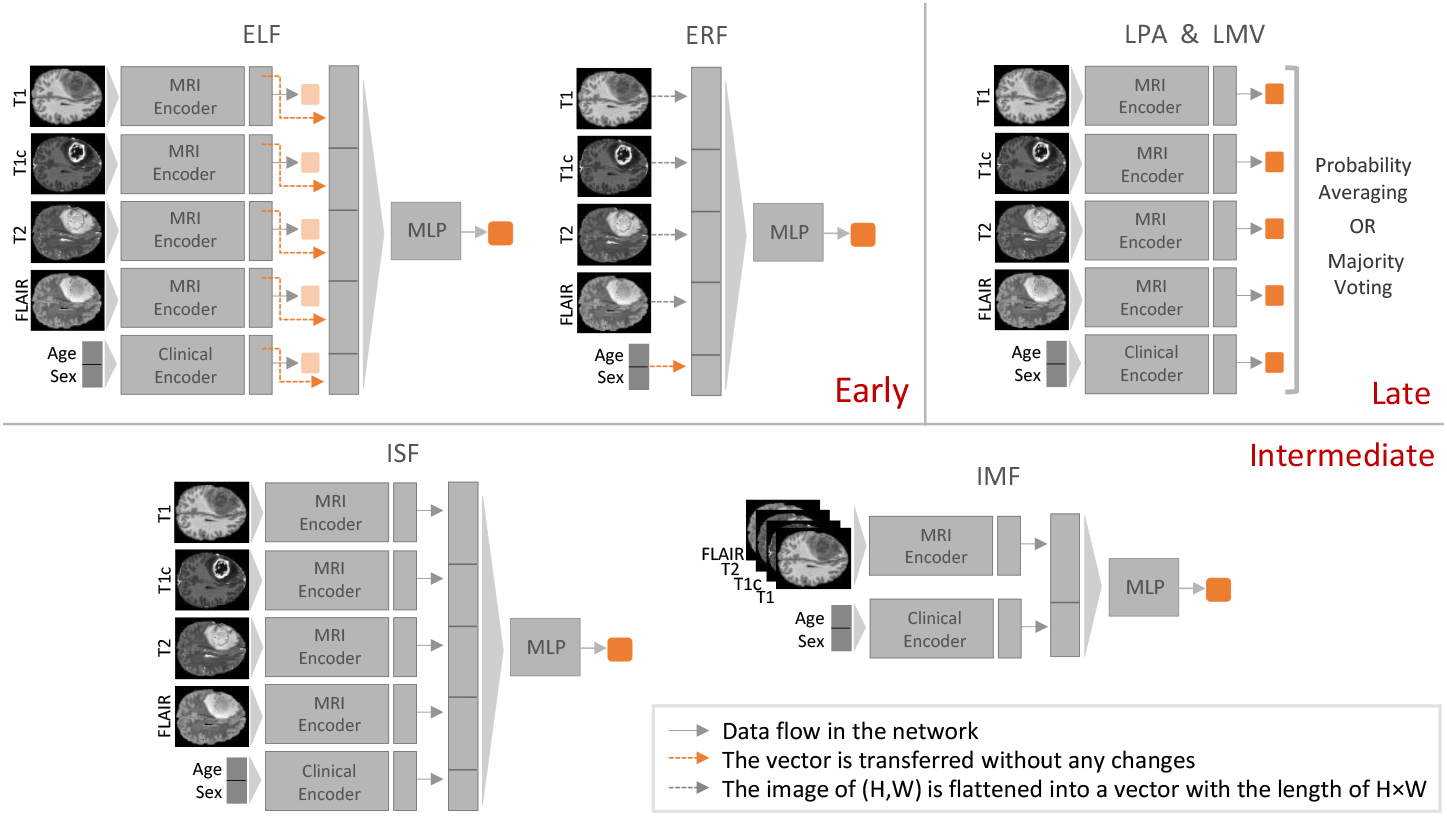
Fusion strategies examined in our framework

In ERF fusion, MRI modalities are flattened into feature vectors and directly concatenated with clinical data to form a single input vector, which is then fed into a multilayer perceptron (MLP)—a common classifier in multimodal learning [1]. In contrast, ELF fusion trains a separate encoder for each modality, concatenates the resulting high-level feature vectors in the final fully connected layers, and inputs them into an MLP classifier.

The ISF fusion strategy uses the same modality-specific encoders as ELF, but fusion occurs at the feature extraction stage. Encoded representations from each modality are concatenated and passed to an MLP classifier identical to that of ELF, allowing interactions between modalities to be captured during the feature learning process. In the IMF, the MRI modalities are provided as separate channels of a single multi-channel input and processed jointly by a shared encoder. The clinical features are fed into an encoder independently, and their embeddings are concatenated with the MRI representation before classification via an MLP. This design enables interactions not only among MRI modalities but also between MRI and clinical data during the feature extraction process.

Finally, in the late fusion strategy, independent models are trained for each modality using the same encoders, and their predictions are aggregated via probability averaging in LPA or majority voting in LMV.

### 3.2 Modality Contribution Scores

To better quantify and interpret the relative contributions of MRI and clinical modalities in glioma grade classification, we use the Modality Weighting Block (MWB) as an alternative to conventional feature concatenation. MWB is an enhanced version of the keyless attention modality fusion architecture of DuKa [21]. Unlike simple concatenation, which treats all modalities equally, MWB learns modality-specific weights prior to fusion, enabling the model to adaptively emphasise more informative inputs. The keyless attention mechanism of MWB avoids the full query–key–value formulation, making it lightweight and efficient. The architecture of MWB is shown in fig. 3. Consider *n* modalities, where each modality *i* (*i* = 1, …, *n*) is represented by an embedding 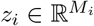, with *M*_*i*_ denoting its dimensionality. Each *z*_*i*_ is first projected into a shared feature space ℝ ^*S*^, yielding *zp*_*i*_ ∈ ℝ^*S*^. A shared MLP with *tanh* as the activation function is applied across all *zp*_*i*_ to compute unnormalised importance scores, which are normalised with softmax to produce contribution scores *{w*_1_, …, *w*_*n*_*}*. The fused representation is obtained as a weighted sum 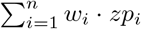, where the weights are learnable parameters optimized jointly with the task objective. By using a shared MLP, MWB enforces direct competition between modalities within the same representational space while keeping the parameter count low. This design can be used for the fusion of features with different dimensions, both enhancing interpretability by yielding explicit modality contribution scores and improving generalization by avoiding over-parameterisation.

**Fig. 3:**
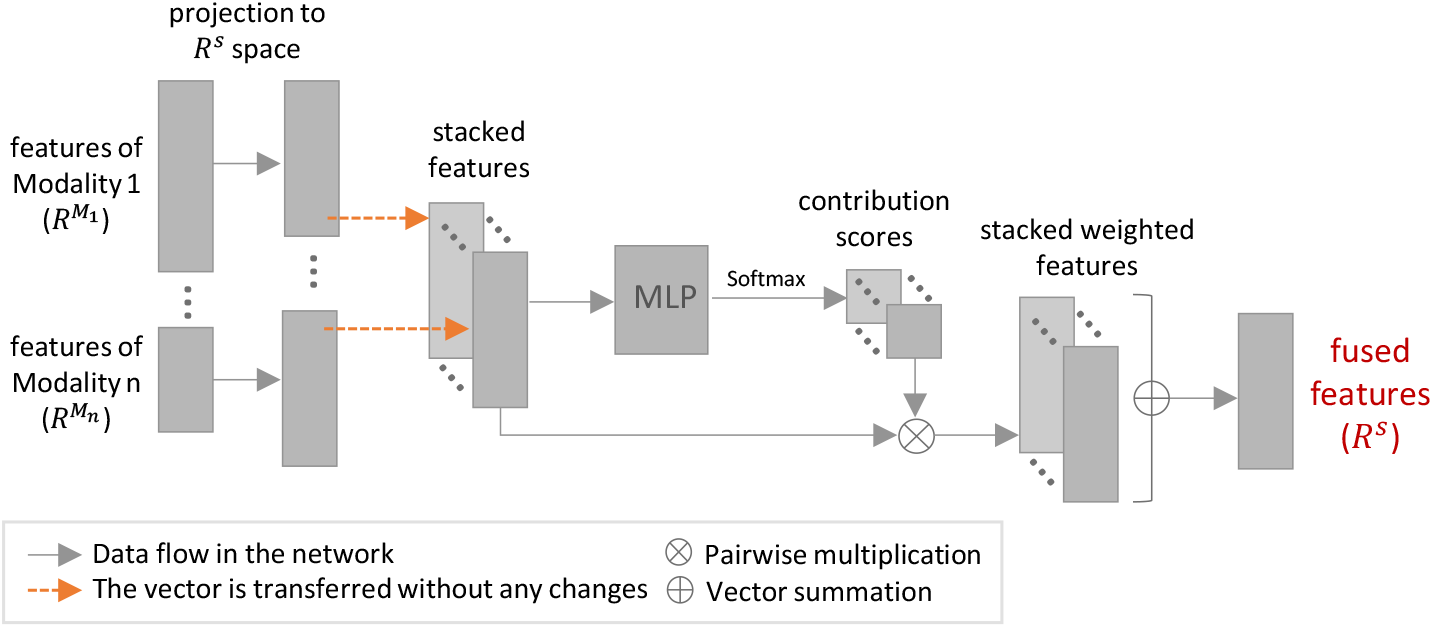
Modality Weighting Block

### 3.3 Dataset Description

We conducted our experiments using the University of California, San Francisco Preoperative Diffuse Glioma MRI (UCSF-PDGM) dataset [22], which includes 501 adult patients with WHO grade 2-4 diffuse gliomas (56, 43, and 402 patients, respectively, for grades 2, 3, and 4) who underwent preoperative MRI and resection between 2015 and 2021. The dataset provides standardized 3T MRI scans across multiple sequences, including T1 (pre- and post-contrast), T2, and FLAIR, along with advanced modalities such as DWI, SWI, and perfusion imaging. It also contains clinical and molecular data (age, sex, tumour grade, IDH mutation, MGMT methylation) and expert tumour segmentations.

### 3.4 Experimental Design

#### Data Preparation

We used data of 495 patients for whom we have both four MRI modalities (T1, T1c, T2, and FLAIR) and clinical features (age and sex). Binary classification (grade 2&3 vs 4) was performed at the slice level, with only tumour-containing slices from each patient included in the dataset (30557 slices). The UCSF-PDGM dataset provides preprocessed MRI images, and we applied modality level min-max normalisation scaling to each slice. Also, min-max normalisation is used for scaling the clinical data. To improve model generalisability, we employed data augmentation consisting of random horizontal and vertical flips, as well as random rotations. To leverage pretrained ImageNet weights in the MRI encoder, all MRI slices were resized to 224×224 pixels.

#### Baseline and Fusion Models

An ImageNet pre-trained DenseNet121 [23] backbone and AutoInt [24] model were selected as the MRI and clinical encoders after a series of preliminary benchmark analysis comparing different architectures, including Swin-Transformers [25] and GBNet [26]. All unimodal baseline models for this work were first trained using DenseNet121 for MRI modalities and AutoInt for clinical features to establish individual modality performance. For MRI inputs, the first convolutional layer of DenseNet121 was adapted to match the number of channels of the corresponding modality. The MLP classifier in the ELF, ISF, and IMF fusion strategies consisted of fully connected layers with sizes [128, 64, 1], whereas the ERF classifier employed a deeper structure of [1024, 512, 256, 128, 1]. Across all models, the weights were initialised with Kaiming initialisation [27]. To ensure a fair comparison of performance, all models were trained on identical dataset splits using the same hardware. All models and fusion strategies were implemented in PyTorch ^3^.

#### Modality Contribution Study

To examine the modality contributions, we first performed an ablation study in which each fusion architecture was trained under two settings: (1) using all MRI modalities together with clinical features, and (2) using MRI modalities only, while keeping all other specifications identical. Second, for the best-performing fusion strategy, we replaced feature concatenation with the proposed Modality Weighting Block. After hyper-parameter tuning, the ablation study was repeated to assess the effect of modality-aware fusion. Modality contribution scores were derived by averaging the contribution scores across all test instances. To ensure robustness, dataset splitting and model training were repeated with three distinct random seeds.

#### Data Splitting and Model Training

We used 5-fold cross-validation, splitting the data at the patient level to prevent data leakage. Each fold consisted of an 80/20 train–test split, with 10% of the training set held out for validation. For the sake of comparison between different fusion strategies, the same folds of the dataset have been used for all the fusion strategies. We trained all models with a batch size of 16 and employed early stopping with a patience of 10 epochs, based on validation loss. The classification objective was optimized using binary cross-entropy with logits loss (BCEWithLogitsLoss), and optimisation was performed using the Adam optimizer, with distinct learning rates assigned to different blocks of layers within architectures. Hyperparameter optimisation was conducted through a combination of Optuna and GridSearch.

#### Evaluation Strategy

To address the class imbalance inherent in the dataset, we employed a sampler-based data loader and evaluated performance using metrics robust to imbalance, including accuracy, weighted F1-score, and Matthews Correlation Coefficient (MCC). The MCC is defined as:

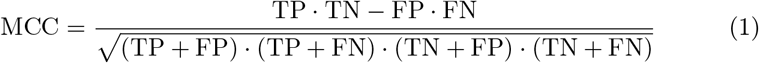

and has some advantages over F1 and accuracy for binary classification [28].

## 4 Results and Discussion

### Baseline Performance

As shown in table 1, unimodal models exhibited clear differences across modalities. T1c achieved the strongest MRI performance (MCC 0.48), whereas T1 was the weakest, and T2 and FLAIR performed similarly. Notably, the clinical model achieved a competitive MCC (0.44) in significantly less training time (0.06 vs. 8 minutes for MRI). These results highlight the importance of modality choice and the robustness and efficiency of clinical features as a valuable complementary modality for MRI in glioma grading. Building on these unimodal baselines, we next investigate the impact of combining MRI and clinical modalities at various fusion levels.

**Table 1:** Performance comparison of Baseline models.

| Metrics | Modality |  |  |  |  |
| --- | --- | --- | --- | --- | --- |
|  | T1 | T1c | T2 | FLAIR | Clinical |
| MCC | 0.34 ± 0.08 | **0.48 ± 0.07** | 0.37 ± 0.07 | 0.37 ± 0.02 | 0.44 ± 0.08 |
| ACC | 0.77 ± 0.03 | **0.83 ± 0.02** | 0.80 ± 0.02 | 0.80 ± 0.02 | 0.80 ± 0.03 |
| F1-weighted (avg) | 0.79 ± 0.03 | **0.84 ± 0.02** | 0.81 ± 0.02 | 0.81 ± 0.02 | 0.82 ± 0.03 |
| Recall-weighted (avg) | 0.77 ± 0.03 | **0.83 ± 0.02** | 0.80 ± 0.02 | 0.80 ± 0.02 | 0.80 ± 0.03 |
| Precision-weighted (avg) | 0.82 ± 0.04 | **0.86 ± 0.02** | 0.82 ± 0.03 | 0.82 ± 0.02 | 0.85 ± 0.03 |
| Training time (minutes) | 8.57 ± 0.79 | 8.70 ± 0.73 | 9.40 ± 1.52 | 8.67 ± 0.44 | **0.06 ± 0.03** |

### Fusion Strategies

In early fusion, ELF achieved the strongest results with MRI alone, showing slight gains in accuracy and recall when clinical data were incorporated, while maintaining consistently high F1 and precision scores. In contrast, ERF underperformed relative to ELF and exhibited a further decline in performance when the clinical modality was added (table 2). These findings highlight two key insights: first, the effectiveness of multimodal learning is highly dependent on the underlying fusion architecture, and second, the inclusion of additional modalities does not inherently guarantee performance improvements, as poorly integrated features may introduce noise or redundancy that hinders model learning and performance. In the late fusion setting (table 3), LMV and LPA demonstrated comparable performance, with the latter showing slightly greater robustness. Incorporating clinical data into MRI inputs improved model performance in both cases, with the most notable gain in MCC observed under LMV. Unlike early fusion, where the addition of clinical data occasionally reduced performance, late fusion consistently benefited from their inclusion. Regarding the intermediate fusion results in table 4, ISF achieved the strongest overall performance, with the inclusion of clinical data yielding consistent improvements across all metrics, particularly MCC and accuracy. This indicates that enabling modality interaction during feature extraction is especially beneficial when combining MRI with clinical features. IMF also benefited from clinical data, with the MCC rising from 0.43 to 0.51; however, overall performance remained slightly below that of ISF. Interestingly, in the intermediate fusion setting, adding clinical modality to MRI data makes all the metric performances more robust.

**Table 2:** Performance comparison of early fusion strategies.

| Metrics | ELF |  | ERF |  |
| --- | --- | --- | --- | --- |
|  | MRI | MRI + Clinical | MRI | MRI + Clinical |
| MCC | **0.48 ± 0.07** | 0.46 ± 0.08 | 0.31 ± 0.09 | 0.28 ± 0.09 |
| ACC | 0.85 ± 0.03 | **0.86 ± 0.03** | 0.79 ± 0.02 | 0.79 ± 0.04 |
| F1-weighted (avg) | **0.85 ± 0.03** | **0.85 ± 0.03** | 0.79 ± 0.02 | 0.79 ± 0.03 |
| Recall-weighted (avg) | 0.85 ± 0.03 | **0.86 ± 0.03** | 0.79 ± 0.02 | 0.79 ± 0.04 |
| Precision-weighted (avg) | **0.85 ± 0.03** | **0.85 ± 0.03** | 0.81 ± 0.03 | 0.80 ± 0.02 |

**Table 3:**
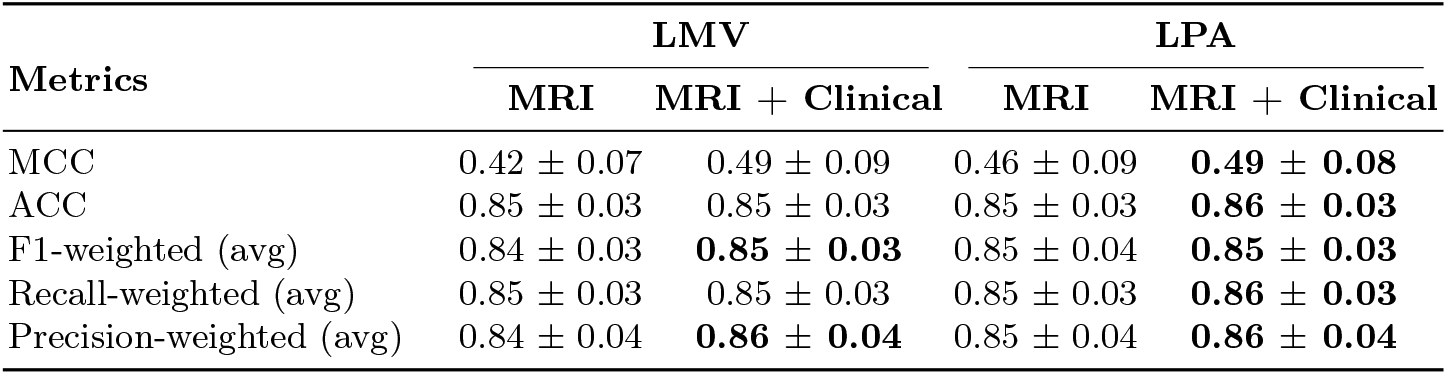
Performance comparison of late fusion strategies.

| Metrics | LMV |  | LPA |  |
| --- | --- | --- | --- | --- |
|  | MRI | MRI + Clinical | MRI | MRI + Clinical |
| MCC | 0.42 ± 0.07 | 0.49 ± 0.09 | 0.46 ± 0.09 | **0.49 ± 0.08** |
| ACC | 0.85 ± 0.03 | 0.85 ± 0.03 | 0.85 ± 0.03 | **0.86 ± 0.03** |
| F1-weighted (avg) | 0.84 ± 0.03 | **0.85 ± 0.03** | 0.85 ± 0.04 | **0.85 ± 0.03** |
| Recall-weighted (avg) | 0.85 ± 0.03 | 0.85 ± 0.03 | 0.85 ± 0.03 | **0.86 ± 0.03** |
| Precision-weighted (avg) | 0.84 ± 0.04 | **0.86 ± 0.04** | 0.85 ± 0.04 | **0.86 ± 0.04** |

**Table 4:** Performance comparison of intermediate fusion strategies.

| Metrics | ISF |  | IMF |  |
| --- | --- | --- | --- | --- |
|  | MRI | MRI + Clinical | MRI | MRI + Clinical |
| MCC | 0.47 ± 0.08 | **0.54 ± 0.07** | 0.43 ± 0.11 | 0.51 ± 0.04 |
| ACC | 0.83 ± 0.05 | **0.86 ± 0.03** | 0.82 ± 0.04 | 0.84 ± 0.02 |
| F1-weighted (avg) | 0.83 ± 0.04 | **0.86 ± 0.03** | 0.83 ± 0.03 | 0.85 ± 0.01 |
| Recall-weighted (avg) | 0.83 ± 0.05 | **0.86 ± 0.03** | 0.82 ± 0.04 | 0.84 ± 0.02 |
| Precision-weighted (avg) | 0.86 ± 0.03 | 0.87 ± 0.03 | 0.85 ± 0.03 | **0.87 ± 0.01** |

### Comparative Analysis Across Fusion Levels

Across all experiments, incorporating clinical data generally improved model performance, except for the early fusion setting. Intermediate fusion strategies consistently outperformed both early and late fusion, with ISF achieving the strongest overall results (MCC = 0.54). While late fusion delivered competitive outcomes when combining MRI and clinical features, its improvements were modest relative to its simplicity. These findings suggest that intermediate fusion enables richer cross-modal interactions by allowing modalities to influence each other during encoding.

A direct comparison between ELF and ISF illustrates this effect. Although both strategies rely on modality-specific encoders followed by an MLP classifier, they differ in how and when integration occurs. ELF combines features only after independent extraction, whereas ISF promotes interaction during encoding, thereby capturing complementary information across modalities. This distinction highlights how the fusion level significantly influences a model’s capacity to harness multimodal synergies. Based on these results, ISF is identified as the most effective fusion strategy for further analysis, balancing predictive performance with clinical interpretability.

An additional insight emerges from the MRI-only experiments. ELF delivered the strongest results among multimodal MRI models, with performance nearly matching the unimodal T1c baseline. In contrast, other multimodal strategies underperformed compared to T1c alone. This emphasizes that multimodality is not inherently superior: the effectiveness of fusion depends on both the choice of modalities and the fusion strategy. Careful design is therefore essential to ensure that multimodal integration enhances rather than dilutes predictive signals.

### Modality Contribution Studies

To further examine modality importance, we replace concatenation in ISF with the proposed Modality Weighting Block.

As shown in table 5, this modification consistently improves performance, with MRI + clinical achieving the highest robust scores across all metrics (MCC = 0.56, accuracy, F1, recall, and precision ≥ 0.87). Compared to concatenation-based ISF, these gains underscore the effectiveness of attention in adaptively weighting modalities rather than treating them equally.

**Table 5:** Performance of the ISF architecture with MWB.

| Metrics | ISF with MWB |  |
| --- | --- | --- |
|  | MRI | MRI + Clinical |
| MCC | 0.51 ± 0.05 | **0.56 ± 0.04** |
| ACC | 0.86 ± 0.03 | **0.87 ± 0.01** |
| F1-weighted (avg) | 0.86 ± 0.02 | **0.87 ± 0.01** |
| Recall-weighted (avg) | 0.86 ± 0.03 | **0.87 ± 0.01** |
| Precision-weighted (avg) | 0.86 ± 0.02 | **0.88 ± 0.01** |

Beyond improved performance, the key strength of this architecture is its ability to quantify the relative importance of each modality. The contribution scores derived from attention weights (fig. 4) reveal that for the MRI-only model, T1c emerges as the most influential modality, while T1, T2, and FLAIR play smaller but complementary roles. When clinical data are included, the attention shifts substantially towards the clinical modalities, which dominate predictive power, particularly for grades 2&3, while the ranking among the scores of different modalities of MRI remains consistent despite their reduced overall contributions.

**Fig. 4:**
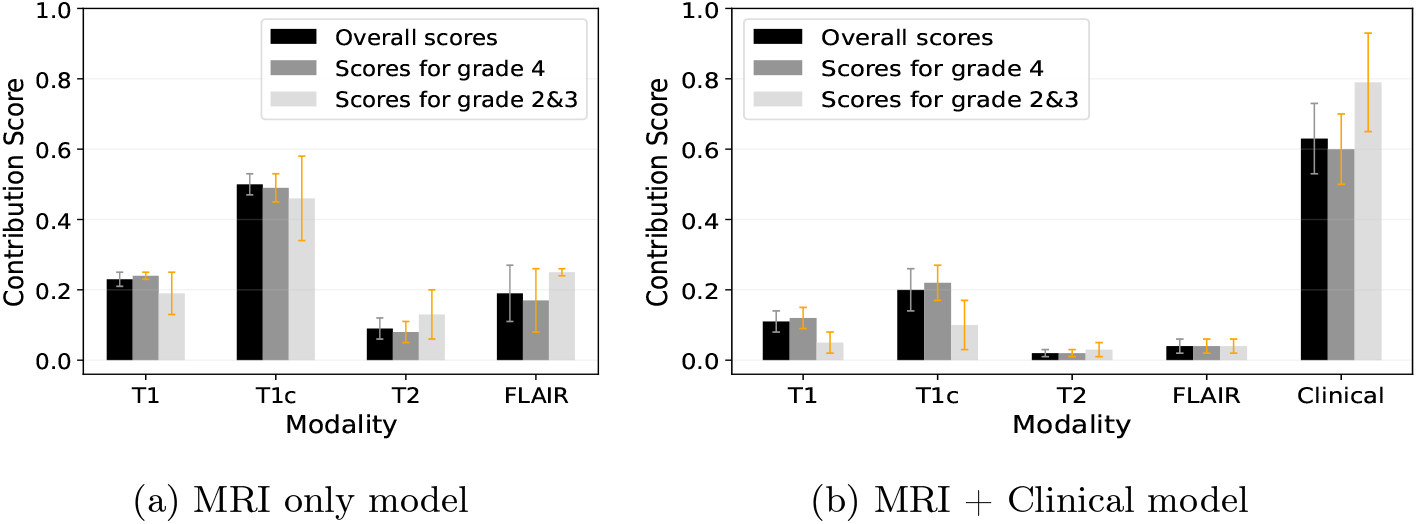
Modality-specific contribution scores of ISF architecture with MWB

These findings demonstrate that modality-aware weighting not only enhances performance and robustness but also provides interpretable insights into how different modalities contribute to outcomes across various glioma tumour grades. The attention-based fusion block allows the model to prioritise informative modalities while down-weighting less relevant signals, resulting in a more effective and transparent multimodal glioma grading model.

## 5 Conclusion

This study presents a multimodal machine learning framework for glioma grading, integrating MRI and clinical data by comparing early, intermediate, and late fusion. Intermediate fusion proved most effective, and adding clinical data generally improved performance. The top-performing ISF model was enhanced with the Modality Weighting Block, which boosted performance and provided interpretable scores. These scores revealed T1c as the key MRI sequence; however, clinical features became dominant when included. The comparative analysis of the fusion strategies in our framework shows that multimodality is not inherently superior—its value lies in the right choice of modalities and fusion strategy, ensuring that integration enhances predictive power instead of diminishing it. While we acknowledge that a key limitation of this work is the small and imbalanced dataset, this research demonstrates that a modality-aware intermediate-fusion approach improves both accuracy and interpretability in glioma grading tasks.

## Data Availability

https://www.cancerimagingarchive.net/collection/ucsf-pdgm/

## Acknowledgments

This work was funded by Taighde Éireann, Research Ireland, through the Research Ireland Centre for Research Training in Machine Learning (grant number 18/CRT/6183) and the Research Ireland Center for Data Analytics (grant number 12/RC/2289 P2)

## Footnotes

3 Code Available: https://github.com/r-kheirbakhsh/Fusion_framework

## References

1. C. Cui, H. Yang, Y. Wang, S. Zhao, Z. Asad, L. A. Coburn, K. T. Wilson, B. A. Landman, and Y. Huo, “Deep multimodal fusion of image and non-image data in disease diagnosis and prognosis: a review,” Progress in Biomedical Engineering, vol. 5, no. 2, 2023.

2. G. Muhammad, F. Alshehri, F. Karray, A. El Saddik, M. Alsulaiman, and T. H. Falk, “A comprehensive survey on multimodal medical signals fusion for smart healthcare systems,” Information Fusion, vol. 76, pp. 355–375, 2021.

3. D. Lahat, T. Adali, and C. Jutten, “Multimodal data fusion: an overview of methods, challenges, and prospects,” Proceedings of the IEEE, vol. 103, no. 9, pp. 1449–1477, 2015.

4. T. Baltrušaitis, C. Ahuja, and L.-P. Morency, “Multimodal machine learning: A survey and taxonomy,” IEEE PAMI, vol. 41, no. 2, pp. 423–443, 2018.

5. Q. T. Ostrom, M. Price, C. Neff, G. Cioffi, K. A. Waite, C. Kruchko, and J. S. Barnholtz-Sloan, “Cbtrus statistical report: primary brain and other central nervous system tumors diagnosed in the united states in 2016—2020,” Neuro-oncology, vol. 25, 2023.

6. Y. Byeon, Y. W. Park, S. Lee, D. Park, H. Shin, K. Han, J. H. Chang, S. H. Kim, S.-K. Lee, S. S. Ahn et al., “Interpretable multimodal transformer for prediction of molecular subtypes and grades in adult-type diffuse gliomas,” NPJ Digital Medicine, vol. 8, no. 1, p. 140, 2025.

7. R. J. Chen, M. Y. Lu, J. Wang, D. F. Williamson, S. J. Rodig, N. I. Lindeman, andF. Mahmood, “Pathomic fusion: an integrated framework for fusing histopathology and genomic features for cancer diagnosis and prognosis,” IEEE Transactions on Medical Imaging, vol. 41, no. 4, pp. 757–770, 2020.

8. P. Guo, L. Li, C. Li, W. Huang, G. Zhao, S. Wang, M. Wang, and Y. Lin, “Multiparametric magnetic resonance imaging information fusion using graph convolutional network for glioma grading,” Journal of Healthcare Engineering, vol. 2022, no. 1, p. 7315665, 2022.

9. T. Usuzaki, K. Takahashi, R. Inamori, Y. Morishita, H. Takagi, T. Shizukuishi,Y. Toyama, M. Abe, M. Ishikuro, T. Obara et al., “Grading diffuse glioma based on 2021 who grade using self-attention-base deep learning architecture: variable vision transformer (vvit),” Biomedical Signal Processing and Control, vol. 91, p. 106001, 2024.

10. X. Long, C. Gan, G. Melo, X. Liu, Y. Li, F. Li, and S. Wen, “Multimodal keyless attention fusion for video classification,” in Proceedings of the aaai conference on artificial intelligence, vol. 32, no. 1, 2018.

11. S. R. Stahlschmidt, B. Ulfenborg, and J. Synnergren, “Multimodal deep learning for biomedical data fusion: a review,” Briefings in bioinformatics, vol. 23, no. 2, p. bbab569, 2022.

12. E. Alpaydin, “Classifying multimodal data,” in The Handbook of Multimodal-Multisensor Interfaces: Signal Processing, Architectures, and Detection of Emotion and Cognition-Volume 2, 2018, pp. 49–69.

13. M. He, K. Han, Y. Zhang, and W. Chen, “Hierarchical-order multimodal interaction fusion network for grading gliomas,” Physics in Medicine & Biology, vol. 66, no. 21, p. 215016, 2021.

14. J. Dhar, N. Zaidi, M. Haghighat, S. Roy, P. Goyal, A. Alavi, and V. Kumar, “Multimodal fusion learning with dual attention for medical imaging,” in WACV, 2025, pp. 4362–4371.

15. J. Wang, L. Yu, and S. Tian, “Cross-attention interaction learning network for multi-model image fusion via transformer,” Engineering Applications of Artificial Intelligence, vol. 139, p. 109583, 2025.

16. L. A. V. Silva and K. Rohr, “Pan-cancer prognosis prediction using multimodal deep learning,” in ISBI, 2020, pp. 568–571.

17. M. Abdelaziz, T. Wang, W. Anwaar, and A. Elazab, “Multi-scale multimodal deep learning framework for alzheimer’s disease diagnosis,” Computers in biology and medicine, vol. 184, p. 109438, 2025.

18. S. Guo, L. Wang, Q. Chen, L. Wang, J. Zhang, and Y. Zhu, “Multimodal mri image decision fusion-based network for glioma classification,” Frontiers in Oncology, vol. 12, p. 819673, 2022.

19. W.-W. Hsu, J.-M. Guo, L. Pei, L.-A. Chiang, Y.-F. Li, J.-C. Hsiao, R. Colen, and P. Liu, “A weakly supervised deep learning-based method for glioma subtype classification using wsi and mpmris,” Scientific Reports, vol. 12, no. 1, p. 6111, 2022.

20. S.-w. Jeong, H.-h. Cho, S. Lee, and H. Park, “Robust multimodal fusion network using adversarial learning for brain tumor grading,” Computer Methods and Programs in Biomedicine, vol. 226, p. 107165, 2022.

21. Z. Liu, X. Wu, Y. Yang, and D. A. Clifton, “Duka: A dual-keyless-attention model for multi-modality ehr data fusion and organ failure prediction,” IEEE Transactions on Biomedical Engineering, vol. 71, no. 4, pp. 1247–1256, 2023.

22. E. Calabrese, J. E. Villanueva-Meyer, J. D. Rudie, A. M. Rauschecker, U. Baid, S. Bakas, S. Cha, J. T. Mongan, and C. P. Hess, “The university of california san francisco preoperative diffuse glioma mri dataset,” Radiology: Artificial Intelligence, vol. 4, no. 6, p. e220058, 2022.

23. G. Huang, Z. Liu, L. Van Der Maaten, and K. Q. Weinberger, “Densely connected convolutional networks,” in Proceedings of the IEEE conference on computer vision and pattern recognition, 2017, pp. 4700–4708.

24. W. Song, C. Shi, Z. Xiao, Z. Duan, Y. Xu, M. Zhang, and J. Tang, “Autoint: Automatic feature interaction learning via self-attentive neural networks,” in CIKM, 2019, pp. 1161–1170.

25. Z. Liu, Y. Lin, Y. Cao, H. Hu, Y. Wei, Z. Zhang, S. Lin, and B. Guo, “Swin Transformer: Hierarchical Vision Transformer using Shifted Windows,” IEEE ICCV, vol. 00, pp. 9992–10002, 2021.

26. M. Horrell, “Gbnet: Gradient boosting packages integrated into pytorch,” Journalof Open Source Software, vol. 10, no. 111, p. 8047, 2025.

27. K. He, X. Zhang, S. Ren, and J. Sun, “Delving deep into rectifiers: Surpassing human-level performance on imagenet classification,” in ICCV, 2015, pp. 1026–1034.

28. D. Chicco and G. Jurman, “The advantages of the matthews correlation coefficient (mcc) over f1 score and accuracy in binary classification evaluation,” BMC genomics, vol. 21, pp. 1–13, 2020.

